# Transplantation of IPSC-Derived Cardiomyocyte Patches for Ischemic Cardiomyopathy

**DOI:** 10.1101/2021.12.27.21268295

**Authors:** Shigeru Miyagawa, Satoshi Kainuma, Takuji Kawamura, Kota Suzuki, Yoshito Ito, Hiroko Iseoka, Emiko Ito, Maki Takeda, Masao Sasai, Noriko Mochizuki-Oda, Tomomi Shimamoto, Yukako Nitta, Hiromi Dohi, Tadashi Watabe, Yasushi Sakata, Koichi Toda, Yoshiki Sawa

## Abstract

**Background:** Despite major therapeutic advances, heart failure remains a life-threatening disorder, with 26 million patients worldwide, causing more deaths than cancer as a non-communicable disease. Therefore, novel strategies for the treatment of heart failure continue to be an important clinical need. Based on preclinical studies, allogenic human-induced pluripotent stem cell-derived cardiomyocyte (hiPSC-CM) patches have been proposed as a potential therapeutic candidate for heart failure. We report the implantation of allogeneic hiPSC-CM patches in a patient with ischemic cardiomyopathy (ClinicalTrials.gov, #jRCT2053190081).

**Methods:** The patches were produced under clinical-grade conditions and displayed cardiogenic phenotypes and safety *in vivo* (severe immunodeficient mice) without any genetic mutations in cancer-related genes. The patches were then implanted via thoracotomy into the left ventricle epicardium of the patient under immunosuppressive agents.

**Results:** Positron emission tomography and computed tomography confirmed the possible efficacy and did not detect tumorigenesis in either the heart or other organs; the clinical symptoms improved 6 months after surgery, without any major adverse events, suggesting that the patches were well-tolerated. Furthermore, changes in the wall motion in the transplanted site were recovered, suggesting a favorable prognosis and the potential tolerance to exercise.

**Conclusions:** This study is the first report of a successful transplant of hiPSC-CMs for severe ischemic cardiomyopathy.

## Introduction

Severe heart failure is a highly lethal disease with high mortality, and patients’ quality of life (QOL) is significantly reduced despite the development of medical treatments^1^. When severe heart failure is reversible, drug treatment is prioritized; however, at the irreversible stage, left ventricular assist device (LVAD) or heart transplantation are first-line therapy. Heart transplantation is an extremely effective treatment for heart failure that can prolong life expectancy. However, there are some drawbacks, such as a shortage of donors worldwide, and securing donors is expected to be increasingly challenging in the future^2^. For LVAD, destination therapy or bridge to transplant is performed, but complications such as infections and cerebral thrombosis are major problems^3^. There is hope for the development of new treatments that can replace artificial hearts and heart transplants for irreversible stages of heart failure and new therapies that prevent worsening heart failure and progression toward the irreversible stage. As an option to overcome such a situation, expectations for regenerative medicine are increasing. In recent years, although cytokine-based angiogenesis treatment using somatic stem cells for heart failure has been performed, it is considered that activation in the hibernating myocardium by angiogenesis does not provide sufficient clinical effects in severe cardiomyopathy that has lost a large number of functional cardiomyocytes. For the above-mentioned diseases, it is necessary to generate cardiomyocytes externally, transplant them into the failing heart, and electrically and functionally integrate them with the recipient heart.

The generation of large numbers of new cardiomyocytes and their integration within recipient hearts is a promising treatment for severely damaged myocardia with few cardiomyocytes. Recently, induced pluripotent stem cells (iPSCs) have been developed from human somatic cells. iPSCs have the property of being able to differentiate into cells of all body organs and are expected to be a source of cell therapy for various diseases ^4,5^. Basic research studies have demonstrated that iPSCs can differentiate into cardiomyocytes as single cells or myocardial tissues^6-10^. These iPSC-derived cardiomyocytes beat spontaneously and produce and release cytokines that induce angiogenesis. Thus, it is conceivable that iPSCs may be used to achieve cardiomyogenesis forming cardiomyocytes that can mechanically contract and repair myocardial tissue via paracrine angiogenic factors.

Here, we report the case of a male patient in his 50s with ischemic cardiomyopathy who had already received maximum anti-heart failure medications such as digitalis, diuretics, ACE inhibitors, angiotensin receptor blockers, beta-blockers, anti-aldosterone drugs, and oral cardiotonics. He was successfully treated with clinical-grade human induced pluripotent stem cell-derived cardiomyocyte (hiPSC-CM) patches with typical cardiogenic phenotypic properties in a first-in-human clinical trial.

## Methods

This study was conducted in accordance with the Declaration of Helsinki. The subject provided informed consent for the use of data. All animal experiments have been approved by the Ethics Committee of Osaka University. The authors declare that all supporting data are available within the article and its Online Data Supplement.

### hiPSC Culture and Cardiomyogenic Differentiation and Purification

The methods for establishing clinical-grade hiPSCs and generation of master cell bank (MCB) of the hiPSC-cell line (QHJI14s04) have been described elsewhere ^11^. Briefly, the clinical-grade iPS cell line (QHJI14s04) was established from the peripheral blood mononuclear cells of a healthy donor (HLA homozygous: HLA-A, HLA-B, HLA-DRB1), showing the most frequent haplotypes in the Japanese population ^12^. QHJI14s04 was generated using episomal plasmids (pCE-hSK, pCE-hUL, pCE-hOCT3/4, pCE-mp53DD, pCXB-EBNA1) and maintained using a feeder-free and xeno-free culture system ^13^ in the cell processing center (Facility for iPS Cell Therapy, CiRA). The evaluation method for ensuring quality and the results are reported separately ^11^. For the production of hiPSC-CMs, the MCB of QHJI14s04 was established under good manufacturing practice conditions.

The QHJI14s04 cells from MCB were cultured on iMatrix511 (Nippi, Tokyo, Japan) coated dishes in Stem Fit Ak03N (Ajinomoto, Tokyo, Japan). Cardiomyogenic differentiation of QHJI14s04 cells was induced using a previously described protocol ^13-15^.

The methods used for evaluating the safety of the manufactured hiPSC-CMs, including assays for general toxicity and determination of *in vitro* and *in vivo* tumorigenicity via cell growth assay, soft agar colony formation assay, whole-genome/whole-exome sequencing analysis, and transplantation of the hiPSC-CMs into severe immunodeficient (NOG) mice, have also been described separately ^11^.

### Cell Patch Preparation

The hiPSC-CM patches were manufactured using temperature-responsive culture dishes ^6^. Prior to cell seeding, the surface of temperature-responsive dishes (UpCell; CellSeed, Japan) was coated with FBS overnight. After freeze-thawing, cells were plated onto the UpCell in DMEM containing 20% FBS and cultured at 37 °C under 5% CO_2_. After 48 h in culture, hiPSC-CM patches were harvested and washed gently with HBSS (+).

### Flow Cytometry

hiPSC-CMs were labeled with anti-cardiac troponin T (cTnT) (REA400, Miltenyi Biotec, Germany) antibodies after fixation with paraformaldehyde. Cell populations, including more than 10000 cells, were resolved using the MACSQuant Analyzer (Miltenyi Biotec). Data were analyzed using the FlowJo (BD Biosciences) software.

### Hematoxylin and Eosin (H&E) and Immunofluorescence Staining

hiPSC-CM patches were fixed in 10% buffered formalin (Fujifilm), paraffin-embedded, and sectioned. Sections were stained with H&E (Muto Pure Chemicals). Immunofluorescence staining was performed with primary antibodies [anti-cTnT (MA5-12960, Neomarkers, Inc, Portsmouth, NH, USA) and anti-connexin 43 (C6219, Sigma-Aldrich, St. Louis, MO, USA)] overnight at 4 °C. Then the sections were labeled with secondary antibodies [Alexa Fluor 488 goat anti-rabbit A11008, and Alexa Flour 555 goat anti-mouse A21422 (both from Thermo Fisher Scientific, Waltham, MA, USA)] at room temperature for an hour. The cell nuclei were stained with Hoechst 33342 (1:100; Dojindo, Kumamoto, Japan), and the preparations were assessed using the confocal laser scanning microscope FV10i (Olympus, Tokyo, Japan). The images were analyzed using the FV10-ASW 3.1 software (Olympus).

### *In vivo* Tumorigenicity Assay

The production lot of the hiPSC-CM used in the clinical trial was subjected to an *in vivo* tumorigenicity test and confirmed to be non-tumorigenic in immunodeficient NOG mice. The detailed procedures of transplantation and histological analysis have been described separately^11^. Mice were euthanized and dissected 16 weeks after transplantation, and the major organs and tissues were carefully observed; gross pathological findings were recorded, and the tissues were stored for histological analysis.

### Patient

A male patient in his 50s was repeatedly admitted to the hospital owing to severe heart failure due to ischemic cardiomyopathy (Figure S1). In 2019, he suffered from acute myocardial infarction and was treated via percutaneous coronary intervention (PCI) targeting the three major coronary arteries. Six months after treatment, he complained of chest pain. A second acute myocardial infarction episode was diagnosed; PCI was introduced via intra-aortic balloon pumping (IABP), and percutaneous cardiopulmonary support was administered due to ventricular fibrillation. However, his condition worsened, and catecholamine and Impella® were then used as therapeutic agents. Although catecholamine treatment and circulatory support could eventually be withdrawn after the 2^nd^ attack (Figure S1), the heart failure persisted (New York Heart Association [NYHA] classification = III). Ultrasonography revealed severe asynergy throughout the wall of the left ventricle (LV) (ejection fraction [EF] = 30%). Positron emission tomography (PET) further revealed ischemia in the middle-apex (anterior wall) and the base-apex (posterior-lateral wall); an infarcted area in the inferior wall was observed despite the absence of significant stenosis in all coronary arteries. The patient was then treated with anti-heart failure medications (maximum dosages), such as beta-blockers, angiotensin-converting-enzyme inhibitors (ACE), and diuretics. Finally, hiPSC-CM transplantation was considered as a therapeutic option.

The regenerative treatment protocol (Japan Registry of Clinical Trials, https://jrct.niph.go.jp/en-latest-detail/jRCT2053190081) using allogeneic iPSC-CM patches was approved by the institutional review boards of Osaka University (#199006-A) and the Ministry of Health, Labor, and Welfare, Japan (#2019-143). The transplantation procedure was performed after obtaining written informed consent from the patient (and his family). The participant consented to have the results of this research work published. The study was conducted in accordance with the principles of the Declaration of Helsinki. During the first 3 months after transplantation, the patient received immunosuppressant drugs, including steroids, tacrolimus hydrate, and mycophenolic acid (mofetil); thereafter, these drugs were discontinued. The cardiac function and myocardial blood perfusion were measured using cardiac computed tomography (CT), wall stress measurements, and myocardial blood flow PET. Tumorigenesis (safety) was investigated via fluorodeoxyglucose-PET (FDG-PET).

### Assessment of Regional Myocardial Displacement Using Four-Dimensional Computed Tomography (4DCT)

The dynamic data from 320-slice cardiac CT images were restored at a constant interval from 10 phases via electrocardiogram synchronization. Subsequently, data complementation (30 phases) using PhyZiodynamics (PhyZiodynamics, 4D motion analysis; Ziosoft Inc., Tokyo, Japan) was performed as reported previously^16, 17^. Next, a 4D motion analysis was performed to evaluate regional myocardial displacement. In the color scale, red indicates a good dynamic region, and the darker the color, the lower the movement.

### End-Systolic Wall Stress (ESS)

Local ESS was calculated as previously reported ^18^ and using the Janz equation ^19^: ESS = P × ΔAc/ΔAw

Where P is the LV end-systolic pressure, and ΔAc and ΔAw are the local cross-sectional areas of the LV cavity and the LV wall at end-systole in each log-axis plane, respectively. The cross-sectional wall area was the area bounded by two lines that are perpendicular to the cavity surface.

In this study, the LV end-systolic pressure estimate was obtained using the following equation ^20^:

P = 0.98 × (systolic blood pressure + (2 × diastolic blood pressure))/3 + 11 mm Hg

Of note, the software can measure local ESS along 30 chords evenly spaced along the epicardial border in each image.

### PET/CT

PET/CT images were acquired using the Biograph Vision 600 (SIEMENS Healthineers, Erlangen, Germany) in the 3-D mode (pixel size: 1.65 mm, slice thickness: 3 mm). FDG-PET scan was performed 60 min after injection of FDG (191 MBq) with glucose loading for heart and whole-body screening. For the evaluation of myocardial blood flow (MBF), a ^13^N-ammonia PET scan was performed immediately after the injection of ^13^N-NH_3_ (approximately 360 MBq) at rest and under stress (120 μg/kg/min of adenosine infusion over 6 min). PET images were reconstructed using the three-dimensional ordered subset expectation maximization algorithm with three iterations/five subsets and Gauss-filtered to a transaxial resolution of 3 mm at full-width at half-maximum (FWHM). Attenuation correction was performed using the unenhanced CT (120 kVp and 200 mAs). MBF was calculated using Cedars (Syngo software; version 5.1; Siemens Healthineers). Myocardial flow reserve was defined as the ratio of MBF at rest to MBF under stress.

## Results

### Properties of hiPSC-CM Patches

The detailed characterization of the hiPSCs and the MCB are presented elsewhere ^11^ and Supplemental Data (Online Table), respectively. The hiPSC-CM patch used in this clinical trial was prepared by a method that cleared the tumorigenicity denial test shown elsewhere^11^. The hiPSC-CM cells used passed all quality inspections, as shown in Table 1.

**Table 1.**
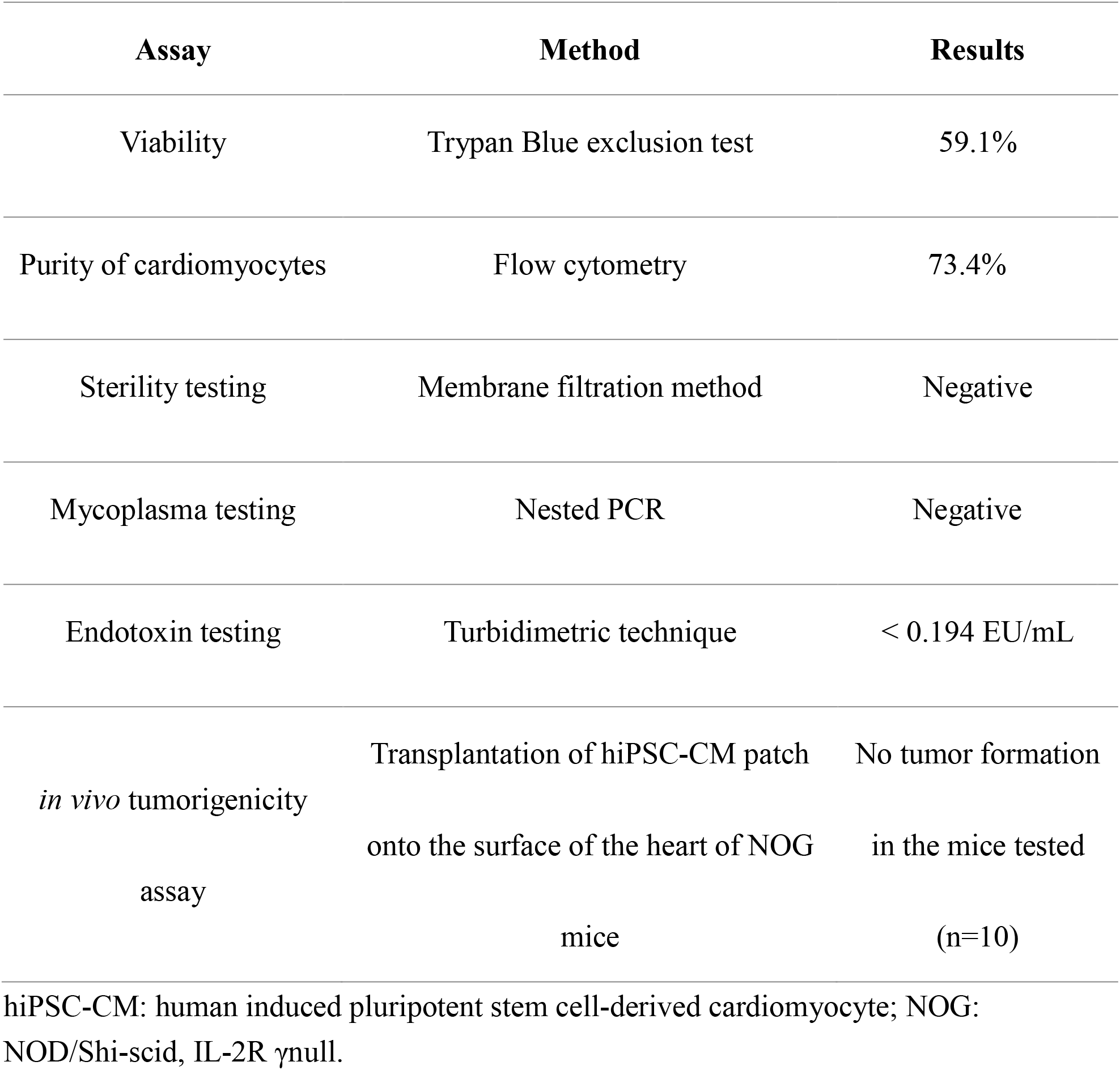
Quality test of hiPSC-CMs.

| Assay | Method | Results |
| --- | --- | --- |
| Viability | Trypan Blue exclusion test | 59.1% |
| Purity of cardiomyocytes | Flow cytometry | 73.4% |
| Sterility testing | Membrane filtration method | Negative |
| Mycoplasma testing | Nested PCR | Negative |
| Endotoxin testing | Turbidimetric technique | < 0.194 EU/mL |
| *in vivo* tumorigenicity assay | Transplantation of hiPSC-CM patch onto the surface of the heart of NOG mice | No tumor formation in the mice tested (n=10) |

hiPSC-CM: human induced pluripotent stem cell-derived cardiomyocyte; NOG: NOD/Shi-scid, IL-2R γnull.

Safety of the patches was assessed *in vivo* using NOG mice; a patch prepared from cells of the same lot as the hiPSC-CMs used in the clinical trial was transplanted into NOG mice and observed for 16 weeks; neither teratomas nor malignant tumors were observed (Table 1, Figure S2). Based on these data, we concluded that the hiPSC-CM patch used in the clinical trial had no risk of tumorigenesis following transplantation into the patient.

hiPSC-CMs were positive for the marker cTnT (Figure 1A, Table 1). Prior to transplantation surgery, hiPSC-CM patches were prepared using temperature-responsive culture dishes (Figure 1B). The immunohistochemistry analysis of the patches revealed well-organized sarcomere structures and a high expression of the gap junction-related protein, connexin 43 (Figure 1C).

**Figure 1.**
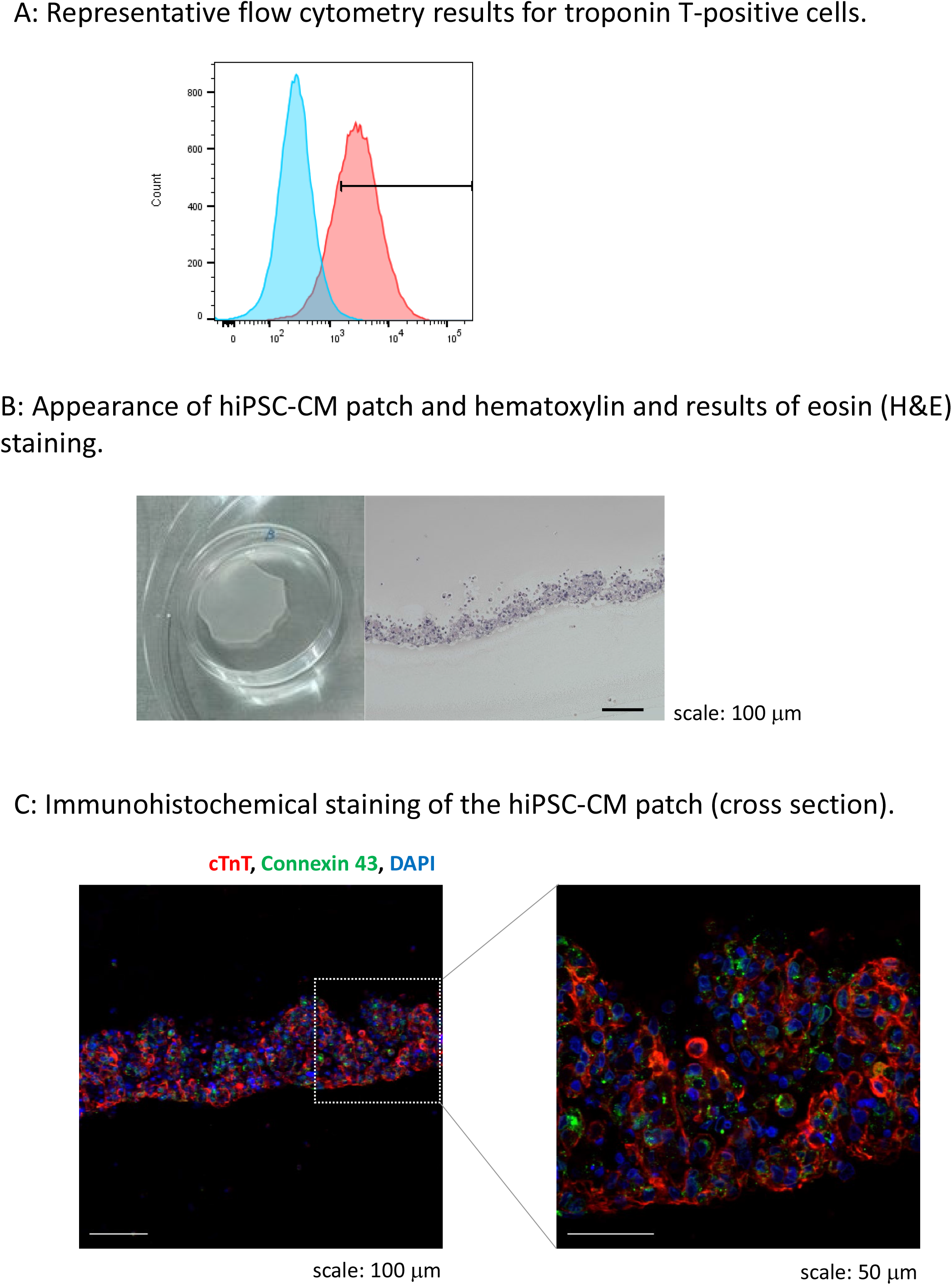
Characteristic properties human induced pluripotent stem cell-derived cardiomyocyte (hiPSC-CM) patch. A: Representative flow cytometry results for troponin T-positive cells (73.4 %) in the hiPSC-CM patch. B: Appearance of hiPSC-CM patch and hematoxylin and results of eosin (H&E) staining. Left: Appearance of an iPSC-CM patch in 60 mm dish; Right: H&E staining of a hiPSC-CM patch (cross-section). Scale bar: 100 μm. C: Immunohistochemical staining of the hiPSC-CM patch (cross-section). Red: cTnT, green: connexin 43, blue: DAPI. Left: Low magnification image, scale bar: 100 μm; Right: High-magnification image, scale bar: 50 μm.

To confirm the absence of neoplastic potential due to critical genomic mutations and survival of foreign genes, whole-genome sequencing analysis was performed on the MCB used in this study. No abnormalities in CNV and SNP were observed for cancer-related genes listed in the Catalogue of Somatic Mutations in Cancer (COSMIC), the Cancer Gene Census ^11^.

To verify whether genomic structural abnormalities are induced during the production process of hiPSC-CM patches, changes in copy number variation (CNV) before and after production were evaluated by array comparative genomic hybridization (CGH) analysis using patch prepared from cells of the same lot as the hiPSC-CMs used in the clinical trial (Figure S3). No CNV abnormality was detected, and it was judged that no significant genomic structural mutation occurred during the manufacturing of the hiPSC-CM patches. CGH analysis showed that the cells produced for this clinical trial were also free of CNV abnormalities.

In order to detect teratoma-forming cells with the hiPSC-MC patch, mRNA expression of Lin28A, a marker of undifferentiated cells, was examined *in vitro* with hiPSC-CMs. As a result, Lin28A expression was below the detection limit (data not shown).

### Clinical Trial

We prepared three allogeneic iPSC-CM patches (3.3 × 10^7^ cells/patch) (Figure 2A). The three patches were successfully transplanted into the epicardium of the LV anterior and lateral walls (Figure 2B) through the fourth intercostal space under IABP support; no other surgical procedures, including coronary artery bypass grafting, were performed. The IABP support was withdrawn the day after surgery, and the catecholamine infusion was discontinued 8 days post-transplantation.

**Figure 2.**
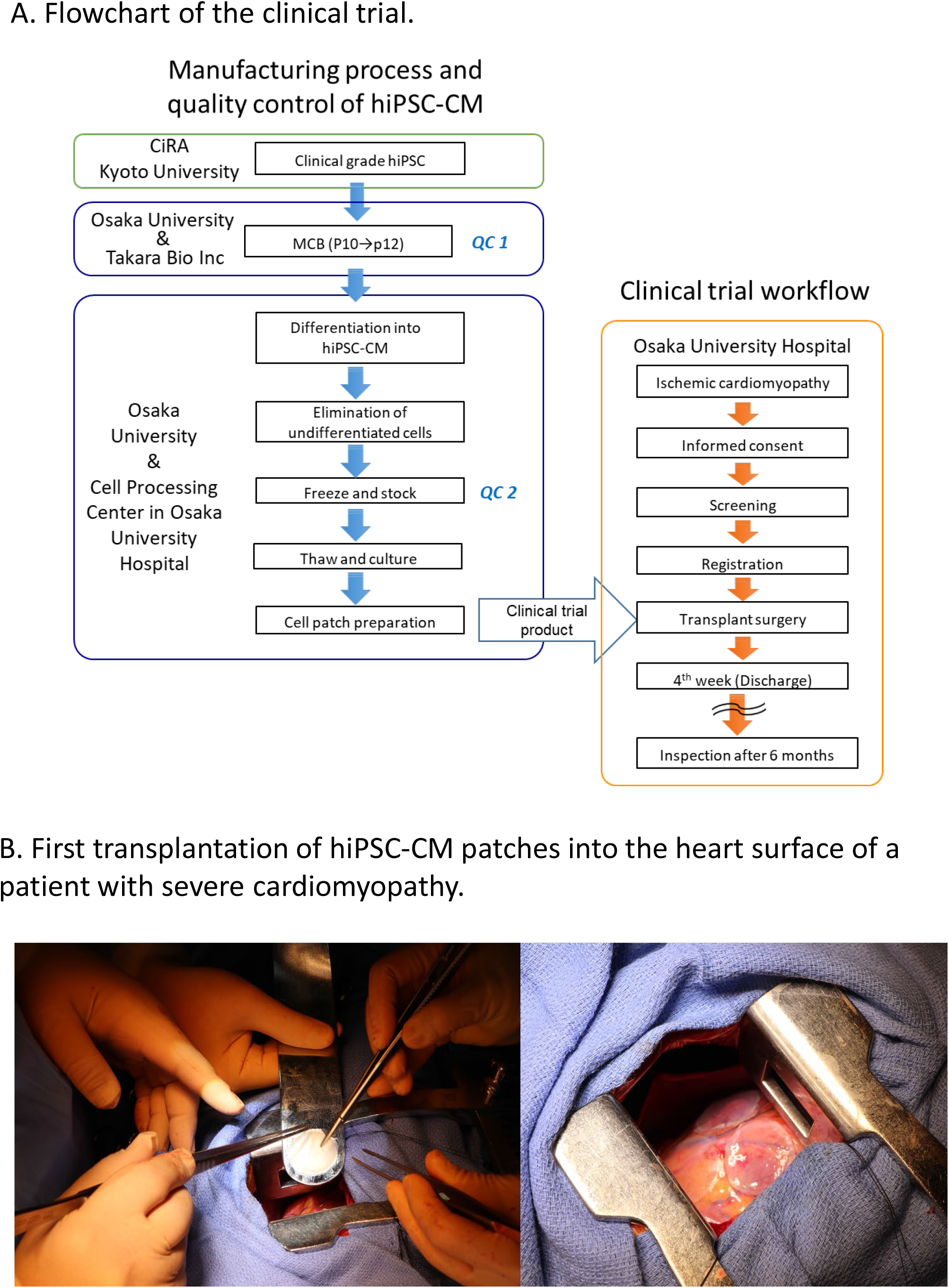
The clinical trial flowchart and first transplantation of human induced pluripotent stem (iPS) cell-derived cardiomyocyte (hiPSC-CM) patches. A: Flowchart of the clinical trial. Clinical-grade iPS cells were established and procured from CiRA (Kyoto University). The hiPSC-CM patches were manufactured in the cell processing center in Osaka University Hospital using the master cell bank (MCB) cells generated in the Center for Gene and Cell Processing of Takara Bio Inc (Kusatsu, Japan). The following quality inspections were performed during the manufacturing process: QC1: Quality check of the MCB (Online Table). Detailed procedure of generation of MCB is described elsewhere ^1^. QC2: viability, purity of cardiomyocytes, sterility, mycoplasma testing, and endotoxin testing (Table 1). B: First transplantation of hiPSC-CM patches into the heart surface of a patient with severe cardiomyopathy. Left: hiPSC-CM patch was transplanted into the epicardium of the anterior and lateral walls of the left ventricle (LV) through the fourth intercostal space under intra-aortic balloon pumping (IABP) support. Right: Three patches were transplanted into the epicardium of the LV anterior and lateral walls.

We did not detect any complications, including arrhythmias, tumor formation, or immunosuppression-related adverse events. Importantly, the patients’ symptoms improved from NYHA III to II at 6 months and 1 year after transplantation (Table 2). Enhanced CT demonstrated that the LV end-systolic volume index (LVESVI) increased after transplantation (Table 2), but was substantially decreased 1 year after transplantation. Peak VO_2_ improved at 6 months and 1 year after transplantation without cardiac rehabilitation (Table 2).

**Table 2.** Changes in cardiac function, exercise tolerance, and heart failure classification after transplant surgery.

| Time point | LVEF<br>(%) | LVESVI<br>(mL/m<sup>2</sup>) | LVEDVI<br>(mL/m<sup>2</sup>) | peak VO<sub>2</sub><br>(mL/min/kg) | NYHA |
| --- | --- | --- | --- | --- | --- |
| pre | 30.0 | 95.71 | 137.42 | 15.4 | 3 |
| 6 month | 28.0 | 113.61 | 158.58 | 18.7 | 2 |
| 1 year | 33.5 | 88.82 | 133.58 | 20.0 | 2 |

LVEF: left ventricular ejection fraction; LVESVI: left ventricular end-systolic volume index; LVEDVI: left ventricular end-diastolic volume index; peak VO<sub>2</sub>: peak oxygen consumption; NYHA: New York Heart Association.

Myocardial displacement was determined based on the regional myocardial wall motion in each region using a dedicated workstation comprising 320 cardiac CT images ^13^. This analysis revealed that the moving distance, especially in the transplantation area, improved significantly at 6 months and 1 year after transplantation (Figure 3), indicating that the myocardium under the transplanted sheets greatly benefited from the angiogenesis mediated by cytokine-paracrine effects. The LV global wall stress, especially that in the LV anterior and lateral walls that received the allogeneic iPSC-CM patches, was reduced after surgery (Figure 4A, B), suggesting the possible attenuation of cardiac fibrosis and the prolongation of survival.

**Figure 3.**
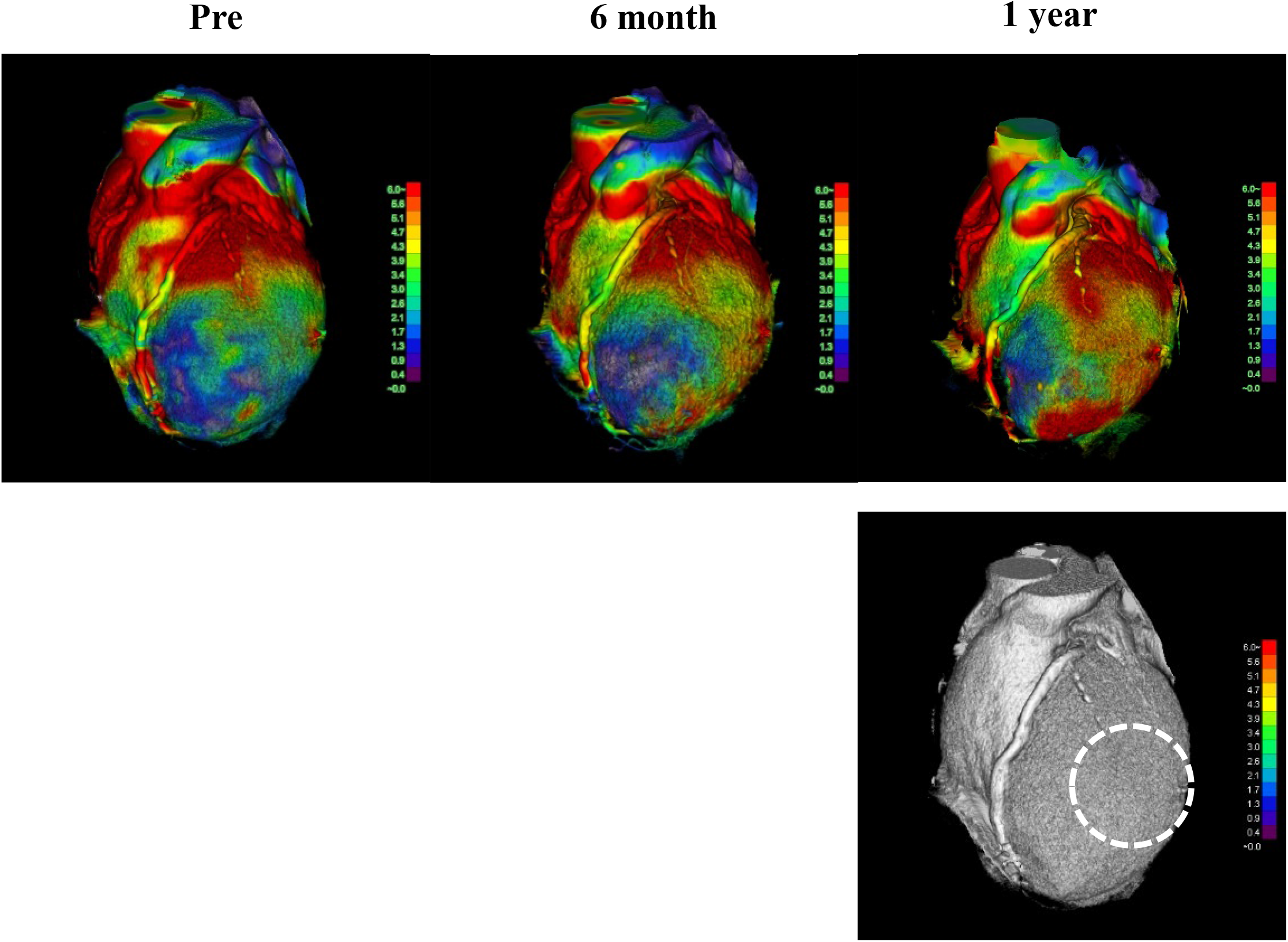
The moving pattern observed via four-dimensional computed tomography (CT). Colors were set so that red indicated a good dynamic area; the darker the color, the lower the movement. The images show the moving pattern in chronological order of the heart at pre-implantation (pre), 6 months, and 1 year after implantation. The dotted line in the bottom figure shows the area where the patches were attached.

**Figure 4.**
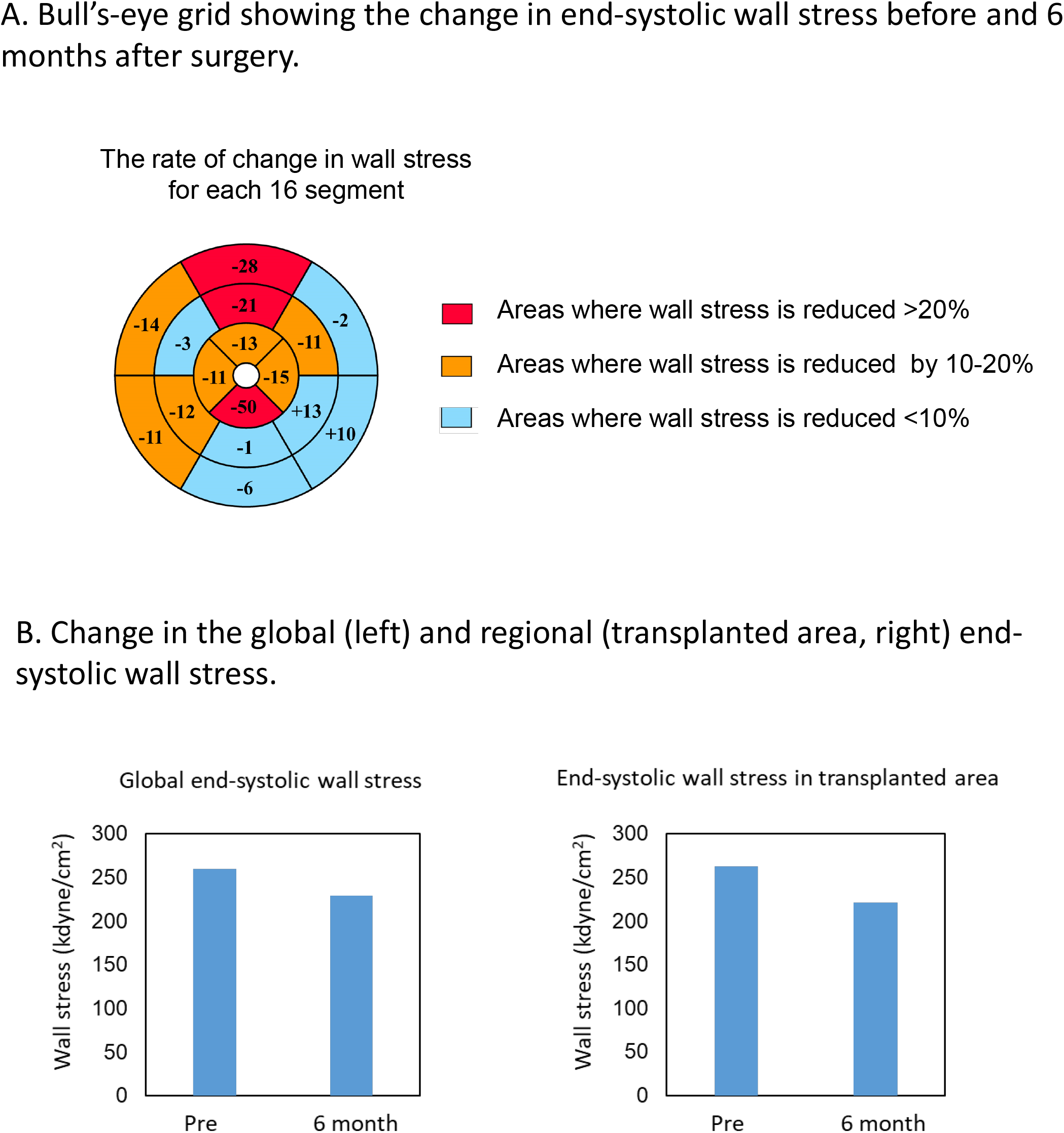
Changes in end-systolic wall stress. A: Bull’s-eye grid showing the changes in end-systolic wall stress before and 6 months after surgery, calculated by computed tomography (CT). Red and orange highlight the areas of reduction in wall stress by over 20% and 10%–20%, respectively. Light blue highlights the remaining areas. B: Change in the global (left) and regional (transplanted area, right) end-systolic wall stress 6 months after transplantation.

According to the ^13^N-ammonia PET images (Figure 5A), the MBF at rest was left anterior descending (LAD) 0.85, circumflex coronary artery (Cx) 0.69, and right coronary artery (RCA) 0.66 ml/g/min at 6 months after transplantation (Figure 5B). However, 1 year after transplantation, the MBF at rest showed a slight decline (LAD 0.65; Cx 0.54; RCA 0.50 ml/g/min) (Figure 5B). The MBF under stress was LAD 1.80, Cx 1.58, and RCA 1.49 ml/g/min 6 months after transplantation but improved 1 year after transplantation (LAD 3.19; Cx 2.93; RCA 3.17 ml/g/min) (Figure 5B). Moreover, from 6 months to 1 year after transplantation, the coronary flow reserves in the whole myocardium greatly improved from 2.16 to 5.30, and the coronary flow reserves in all segmental regions of LV were also dramatically ameliorated (6 months vs. 1 year: LAD 2.12 vs. 4.90; Cx 2.27 vs. 5.40; RCA 2.21 vs. 6.47) (Figure 5C). Furthermore, changes in LV wall motion in the transplanted site were quite dramatic comparing the values at rest with those under stress conditions, and the coronary flow reserve values were similar to those in healthy individuals^14^. Overall, this result suggests a favorable prognosis^15^ and the potential tolerance to exercise. Notably, no medications were introduced or removed post-transplantation. Consistent with the potential tumorigenic safety reported in the preclinical study, the FDG-PET analysis revealed no tumorigenesis after transplantation (Figure 6).

**Figure 5.**
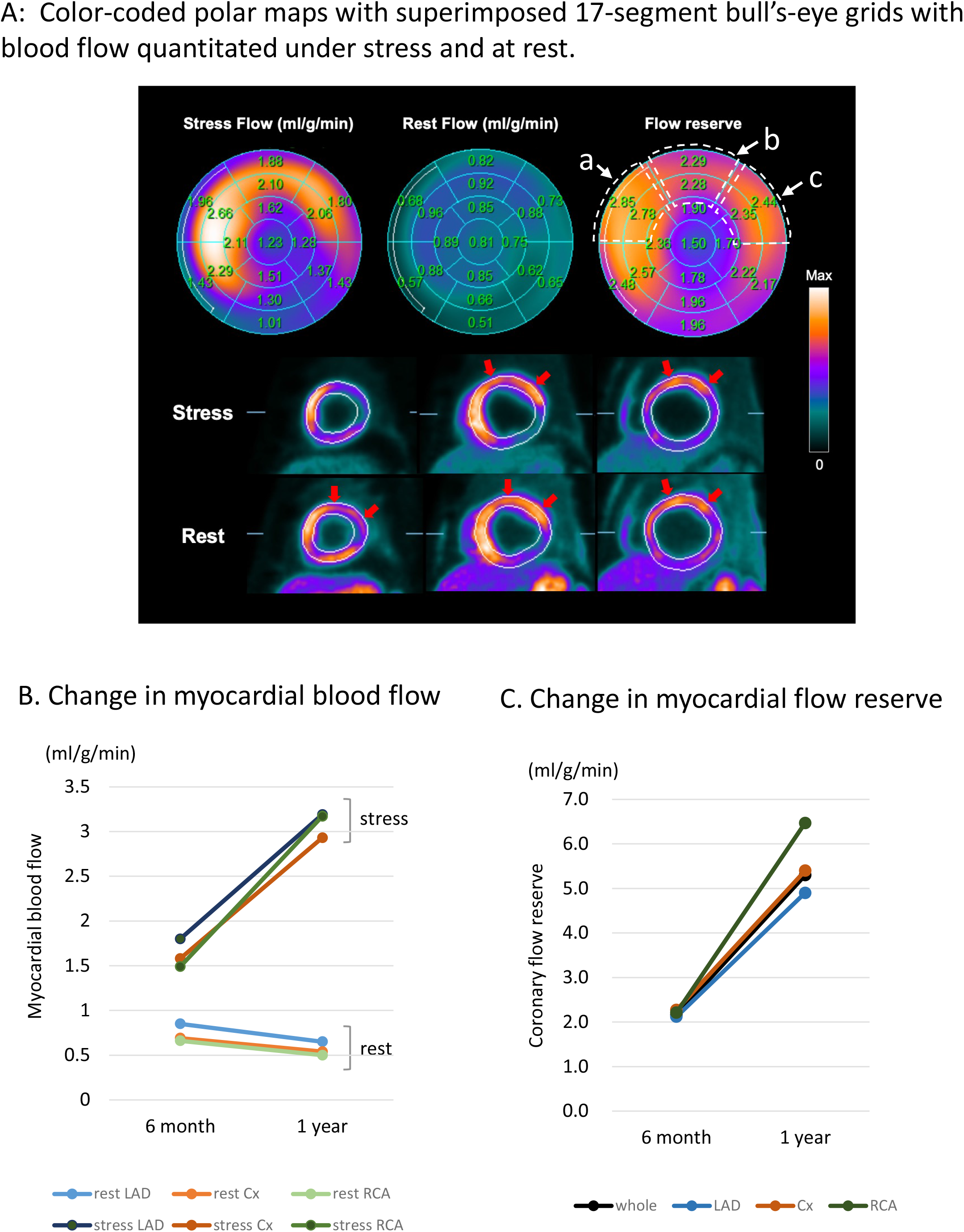
^13^N-ammonia positron emission tomography (^13^N-ammonia PET) images of myocardial blood flow at rest and during stress after human induced pluripotent stem cell-derived cardiomyocyte (hiPSC-CM) sheet implantation. A. Color-coded polar maps with superimposed 17-segment bull’s-eye grids with blood flow quantitated under stress and at rest, as well as flow reserve values, are shown in each segment (upper figures). Myocardial blood flow images (short axis) are shown in the lower figures. Red arrows indicate preserved myocardial flow reserves in the anteroseptal to lateral segments. The areas surrounded by the dotted line in the flow reserve figure show a: anteroseptal, b: anterior, and c: anterolateral regions, and the average values of the two area parts were calculated. B: Change in myocardial blood flow. Myocardial blood flow in the segmental regions LAD, Cx, and RCA at rest and under stress. LAD; left anterior descending coronary artery, Cx; circumflex coronary artery, RCA; Right coronary artery. C: Change in myocardial flow reserve. Myocardial flow reserve in whole and each segmental region at 6 months and 1 year after hiPSC-CM patch transplantation.

**Figure 6.**
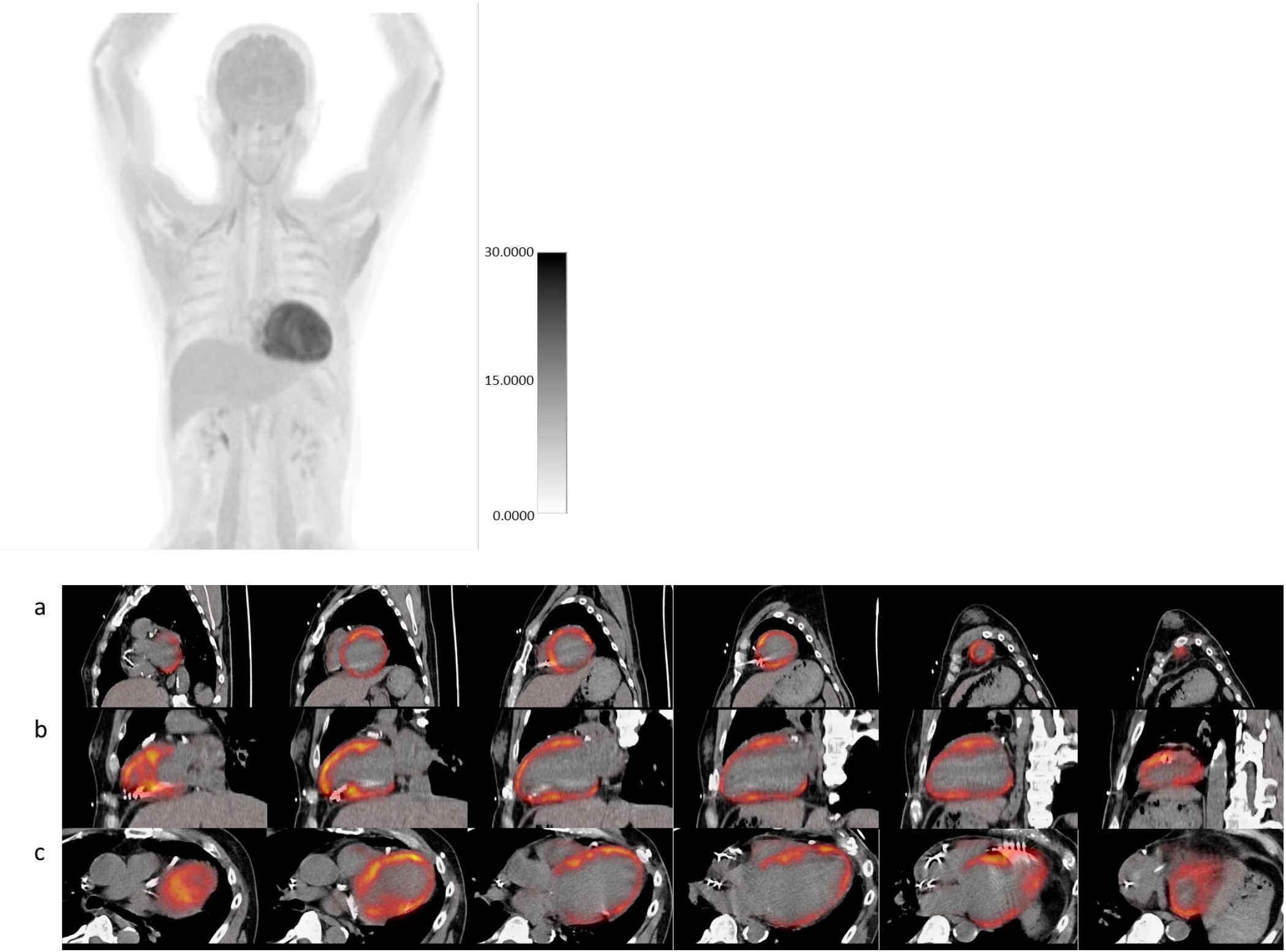
Fluorodeoxyglucose-positron emission tomography (FDG-PET) of the whole-body and the whole heart. FDG-PET showed no obvious abnormal accumulation in the whole-body (top panel: maximum intensity projection) and the whole heart (bottom panel: PET/computed tomography (CT) fusion of short axis (a), the long vertical axis (b), and long horizontal axis (c) views), indicating no tumorigenesis after the transplantation of human induced pluripotent stem cell-derived cardiomyocytes.

## Discussion

Here, we report the first-in-human trial of clinical-grade hiPSC-CM patches to treat ischemic cardiomyopathy in a patient and describe its potential efficacy and safety.

Shiba et al. ^21^ reported that arrhythmias markedly increased in the early phase after transplantation of hiPSC-CMs using a needle. By contrast, we did not detect lethal arrhythmias or tumorigenesis after transplantation in the clinical case. Moreover, our preclinical data^8^ also revealed that the clinical-grade hiPSC-CM patches are non-tumorigenic and non-arrhythmogenic and might be a safe methodology to deliver cardiac cells. Therefore, we assume that the reported arrhythmias did not occur due to transplantation of cardiomyocytes, but rather due to the cell introduction method, particularly due to the needle injection; nonetheless, further clinical evaluations are needed for safety assurance^21,22^.

The major question in the context of hiPSC-CM patches is whether the transplanted cardiac tissues may be electrically integrated within the recipient’s heart, i.e., whether they can undergo “cardiomyogenesis” in a damaged heart with few functional cardiomyocytes. Transplanted cardiomyocyte patches have been reported to undergo contraction/relaxation^23^ or repeated electrical potential activation^24^ in the recipient heart in a synchronous fashion. The cardiogenic properties (both histological and functional) of cardiomyocyte patches were also demonstrated in our preclinical study^11^ (Figure 1). Importantly, in the preclinical study, we observed that the transplanted cardiac tissues facilitated functional recovery; however, the extent of the mechanical contribution of the transplanted cardiomyocytes to the contractile force of the diseased heart needs further investigation.

The functional recovery may also depend on angiogenesis, which positively impacts the hibernating myocardium^25, 26^. In particular, the generation of new functional blood vessels, as we detected in the preclinical experiments^11^, may be sufficient to allow the effective perfusion of blood to hibernating myocytes and to improve the coronary flow reserve in damaged hearts. Based on the *in vitro* cytokine expression profile, the angiopoietin family may have a role in the maturation of blood vessels^11^. The decrease in the resistance of the peripheral coronary arteries may contribute to the improvement of the global coronary flow reserve in the ischemic myocardial tissue and may lead to tolerance to exercise.

In this study, PET detected improved LV wall motion upon transplantation of the patches, especially under stress, accompanied with ameliorated coronary flow reserve, suggesting a potential improvement in exercise tolerance. Furthermore, our PET study revealed that MBF at rest gradually decreased, suggesting that the myocardium could function under low blood flow at rest. The damaged myocardium showed time-course recovery after transplantation.

In this clinical trial, we suspended the administration of immunosuppressant drugs 3 months after transplantation. Notably, we could not clearly elucidate whether transplanted patches survived in the human heart. However, we speculate that the changes in cardiac function, as well as the preserved coronary flow reserve, were mainly dependent on the angiogenic action, which was sufficient to achieve the clinical needs in the treatment of severe heart failure. Further research is warranted to increase the effectiveness of hiPSC transplantation therapy. For instance, a method of monitoring cell survival after transplantation, a detailed analysis of the immune response and immune control mechanism to establish a more appropriate immunosuppressive agent administration routine, and attempts to co-transplant hiPSCs with other types of cells^27^ or tissues^28^ to enhance cardiomyogenesis should be considered.

In conclusion, our clinical-grade hiPSC-CM patches may show great promise for the treatment of ischemic cardiomyopathy and appear to be safe and effective. Although we successfully transplanted hiPSC-CM patches to a patient with ischemic cardiomyopathy without any serious adverse events, further clinical evaluations in a higher number of patients with ischemic heart failure are needed to ascertain the feasibility, safety, and efficacy of this methodology.

## Data Availability

All data produced in the present study are available upon reasonable request to the authors

## Non-standard Abbreviations and Acronyms

4DCT: four-dimensional computed tomography
CGH: comparative genomic hybridization
CiRA: Center for iPS Cell Research and Application, Kyoto University
CNV: copy number variation
Cx: circumflex coronary artery
cTNT: cardiac Troponin T
ESS: end-systolic wall stress
FDG-PET: fluorodeoxyglucose-positron emission tomography
FWHM: full-width at half-maximum
HBSS(+): Hanks’ Balanced Salt Solution
H&E: hematoxylin and eosin
hiPSC-CM: human induced pluripotent stem cell-derived cardiomyocyte
IABP: intra-aortic balloon pumping
LAD: left anterior descending coronary artery
LV: left ventricle
LVESVI: left ventricle end-systolic volume index
MBF: myocardial blood flow
MCB: master cell bank
MI: myocardial infarction
NOG: NOD/Shi-scid, IL-2R γnull
NYHA: New York Heart Association classification
PCI: percutaneous coronary intervention
peak VO_2_: peak oxygen uptake
PET: positron emission tomography
RCA: right coronary artery

## Acknowledgments

We thank the staff of the Department of Cardiovascular Surgery and the Department of Frontier Regenerative Medicine for manufacturing clinical trial products. We also thank the clinical staff in our institution for patient care and data collection.

## Clinical Trial Registration Details

Trial ID: jRCT2053190081; “Clinical trial of human (allogeneic) iPS cell-derived cardiomyocytes patch for ischemic cardiomyopathy” (https://jrct.niph.go.jp/en-latest-detail/jRCT2053190081).

## Data Availability

Most data supporting the findings of this study are available within the article and Supplemental Material or from the corresponding author upon reasonable request. Some of the data are not publicly available due to them containing information that could compromise donor privacy.

## Supplemental Material

Online Data: Figures S1–S4; Table S1.

