## Supplemental Figure S1, S2, S3 and Table S1 for "Transplantation of IPSC-Derived Cardiomyocyte Patches for Ischemic Cardiomyopathy"

**Short Title:** Translational research of iPSC for ICM


**Authors:**Shigeru Miyagawa, M.D., Ph.D.[1]*, Satoshi Kainuma, M.D., Ph.D.[1], Takuji Kawamura, M.D., Ph.D.[1], Kota Suzuki, M.D.[1], Yoshito Ito, M.D.[1], Hiroko Iseoka, Ph.D.[1], Emiko Ito, Ph.D.[1], Maki Takeda, Ph.D.[1], Masao Sasai, Ph.D.[1], Noriko Mochizuki-Oda, Ph.D.[1], Tomomi Shimamoto, B.S.[1], Yukako Nitta, B.S.[1], Hiromi Dohi, Ph.D.[2], Tadashi Watabe, M.D., Ph.D.[3], Yasushi Sakata, M.D., Ph.D.[4], Koichi Toda M.D., Ph.D.[1], Yoshiki Sawa, M.D., Ph.D.[1]

**Affiliations:**

[1]Division of Cardiovascular Surgery, Department of Surgery, Osaka University Graduate School of Medicine, Suita, Osaka, Japan

[2]Center for iPS Cell Research and Application, Kyoto University, Japan

[3]Department of Nuclear Medicine and Tracer Kinetics, Japan

[4]Department of Cardiology, Osaka University Graduate School of Medicine, Suita, Osaka, Japan.

*** Address Correspondence to:**

Shigeru Miyagawa, M.D., Ph.D.

Division of Cardiovascular Surgery, Department of Surgery, Osaka University Graduate School of Medicine, 2-2, Yamada-oka, Suita, Osaka 565-0871, Japan

ORCID iD: 0000-0003-0015-6569

**Online Data**

**Figure S1.** Timeline of symptoms and treatment

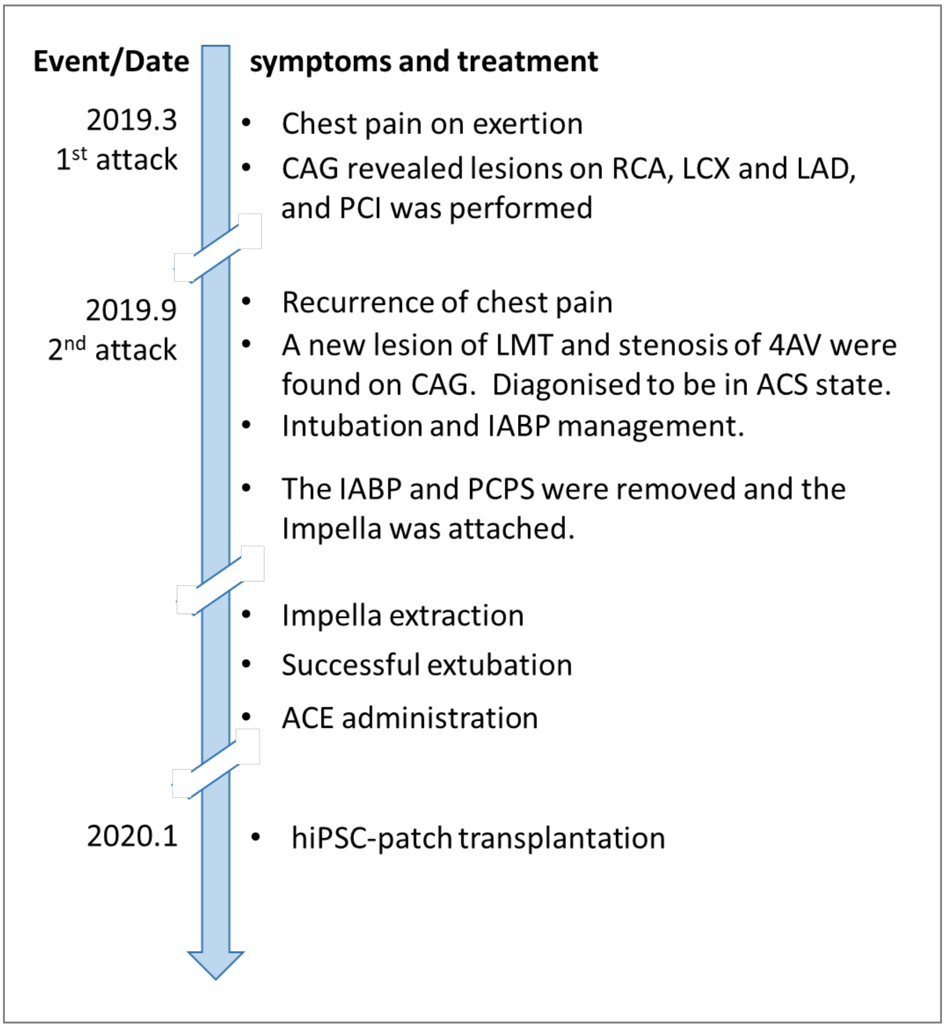


**Figure S1.** Timeline of symptoms and treatment.

The date in parentheses indicates the number of days since the 1st attack. CAG indicates coronary angiography; RCA, right coronary artery; LCX, left circumflex coronary artery branch; LAD, left anterior descending coronary artery; PCI, percutaneous coronary intervention; LMT, left main coronary trunk; AV, atrioventricular node branch; ACS, acute coronary syndrome; IABP, intra-aortic balloon pumping; PCPS, percutaneous cardiopulmonary support; and ACE, angiotensin-converting enzyme inhibitor.

**Figure S2.** Results of histochemical analysis of *in vivo* tumorigenicity test

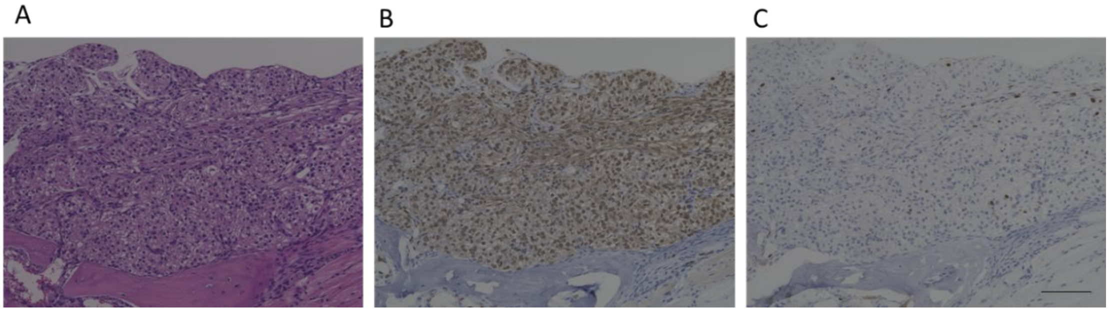


**Figure S2.** Representative images of H&E staining (A), immunostaining of Lamin (B), and Ki67 (C). Scale bar: 100 µm

No histological findings suggestive of tumorigenicity were found.

**Figure S3.** Array CGH analysis

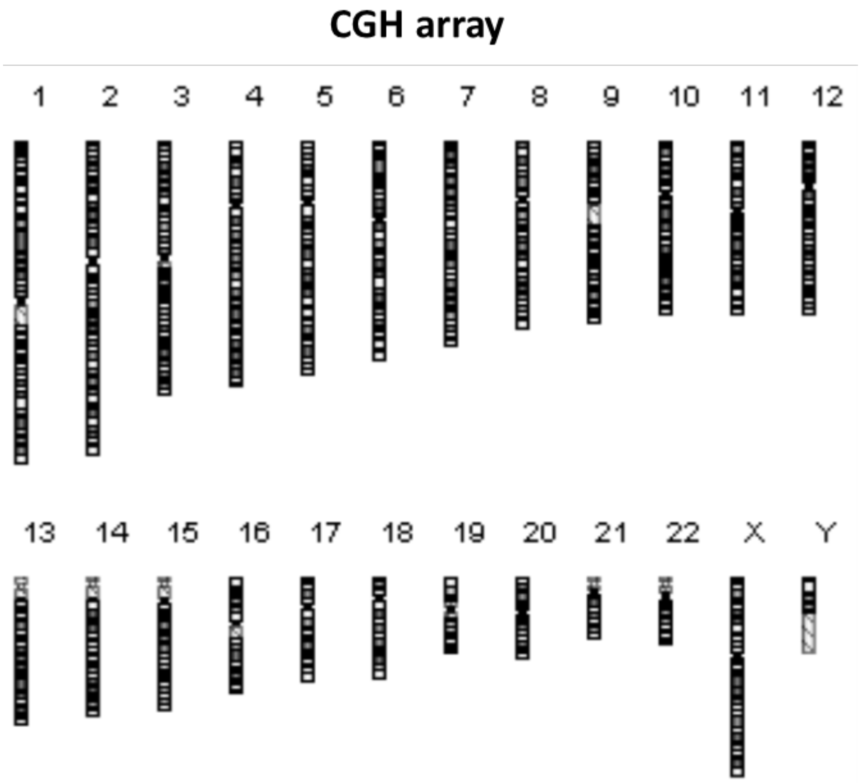


**Figure S3.** Array CGH analysis

Genomic DNA was extracted from cells. After confirming its quality, genomic DNA was fragmented and fluorescently labeled using the SureTag DNA Labeling Kit. Undifferentiated iPSCs were used as controls. Competitive hybridization was performed using SurePrint G3 Human CGH Microarray, 2× 400K with the specified combination, and copy number change analysis was performed.

No CNV was observed in the hiPS-CM patch. If CNV was observed, dots (red for amplification and green for deletion) were displayed at that position.

**Table S1**: Quality test of master cell bank (MCB)

| Assay | Method | Criteria | Results |
|---|---|---|---|
| Sterility Test | Direct inoculation method | Negative | Negative |
| Endotoxin test | Kinetic turbidimetric test | Negative | Negative |
| Mycoplasma Test | DNA staining method and direct culture method | Negative | Negative |
| 200 Median cell profiles | Examination by transmission electron microscopy to detect viruses, virus-like particles, or extraneous agents, including mycoplasmas, yeasts, fungi, or bacteria | Negative | Negative |
| Detection of Reverse transcriptase enzymatic activity assay | Real-time fluorescent product enhanced reverse transcriptase (F-PERT) assay | Negative | Negative |
| Detection of human viral pathogens | Real-Time PCR (*HIV1* and *2* proviruses, *HAV*, *HBV*, *HCV*, *HHV-6*, *HHV-7*, *HHV-8*, *hCMV*, *EBV*, *SV40*, and *B19*) | Negative | Negative |

| | | | |
|---|---|---|---|
| *In vitro* virus assay | Inoculated into MRC-5, Vero C1008, and HeLa cell cultures and observed for the presence of cytopathology or hemadsorbing virus contamination | Negative | Negative |
| *In vivo* virus assay | Inoculated into adult mice, suckling mice, guinea pigs, and embryonated eggs, and observed the indications of any adventitious agents | Negative | Negative |
| Retrovirus assays | Co-cultivation assay using F-PERT | Negative | Negative |
| STR genotyping | PCR-capillary electrophoresis | Match with the original iPS cell profile | Match with the original iPS cell profile |

STR, Short Tandem Repeat.
